# Cortical abnormalities and identification for first-episode schizophrenia via high-resolution magnetic resonance imaging

**DOI:** 10.1101/2020.02.05.20020768

**Authors:** Lin Liu, Long-Biao Cui, Xu-Sha Wu, Ning-Bo Fei, Zi-Liang Xu, Di Wu, Yi-Bin Xi, Peng Huang, Karen M. von Deneen, Shun Qi, Ya-Hong Zhang, Hua-Ning Wang, Hong Yin, Wei Qin

## Abstract

**Intro:** Evidence from neuroimaging has implicated abnormal cerebral cortical patterns in schizophrenia. Application of machine learning techniques is required for identifying structural signature reflecting neurobiological substrates of schizophrenia at the individual level. We aimed to detect and develop a method for potential marker to identify schizophrenia via the features of cerebral cortex using high-resolution magnetic resonance imaging (MRI).

**Method:** In this study, cortical features were measured, including volumetric (cortical thickness, surface area, and gray matter volume) and geometric (mean curvature, metric distortion, and sulcal depth) features. Patients with first-episode schizophrenia (n = 52) and healthy controls (n = 66) were included from the Department of Psychiatry at Xijing Hospital. Multivariate computation was used to examine the abnormalities of cortical features in schizophrenia. Features were selected by least absolute shrinkage and selection operator (LASSO) method. The diagnostic capacity of multi-dimensional neuroanatomical patterns-based classification was evaluated based on diagnostic tests.

**Results:** Mean curvature (left insula and left inferior frontal gyrus), cortical thickness (left fusiform gyrus), and metric distortion (left cuneus and right superior temporal gyrus) revealed both group differences and diagnostic capacity. Area under receiver operating characteristic curve was 0.88, and the sensitivity, specificity, and accuracy of were 94%, 82%, and 88%, respectively. Confirming these findings, similar results were observed in the independent validation. There was a positive association between index score derived from the multi-dimensional patterns and the severity of symptoms (*r* = 0.40, *P* < .01) for patients.

**Discussion:** Our findings demonstrate a view of cortical differences with capacity to discriminate between patients with schizophrenia and healthy population. Structural neuroimaging-based measurements hold great promise of paving the road for their clinical utility in schizophrenia.

## 1. Introduction

Schizophrenia is regarded as a severe mental disorder that has a profound effect on both human health and society (Owen, Sawa, & Mortensen, 2016). “Living with schizophrenia,” in the words of Trevor Turner, “remains hard work,” as Barnett recently mentioned (Barnett, 2018). Over the last decade, in addition to burgeoning evidence showing that schizophrenia is characterized by dramatic structural alterations in the brain, significant variability of brain structures implies that there is overwhelming biological heterogeneity in schizophrenia (Brugger & Howes, 2017). Identifying the determinants of neuroanatomical differences as distinguishing features seems to be a promising step in understanding the nature of schizophrenia and facilitating its diagnosis in the psychiatric domain.

It is highly plausible to link cortical neuroanatomical features to schizophrenia. A recent study based on 4474 patients with schizophrenia and 5098 control subjects presents widespread cortical neuroanatomical abnormalities most prominently in frontal and temporal lobe regions (van Erp et al., 2018). Previous magnetic resonance imaging (MRI) studies have reported differences between patients with schizophrenia and healthy controls (HCs) in the features of cerebral cortex such as cortical thickness, surface areas, gray matter volumes, sulcal depth, metric distortion, and mean curvature (Csernansky et al., 2008; Lyall et al., 2019; Schultz et al., 2013; Wisco et al., 2007; Xiao et al., 2015; Xie et al., 2019; Xu et al., 2017). However, most studies used a mass-univariate method, and these cortical features mentioned above were generally explored in isolation. Notably, these morphological parameters have been used for classifying patients with amnestic mild cognitive impairment via a multivariate method (S. Li et al., 2014). The multivariate method that is superior to the mass-univariate method, which enables treatment of all cortical features together and allows us to examine the relationships among individual features beyond their own value. Thus, this approach could provide valuable insights into the multifactorial etiology of brain disorders. The multivariate method has previously been applied in some brain disorders, e.g., autism spectrum disorder (Ecker et al., 2010), multiple sclerosis (Bendfeldt et al., 2012), and schizophrenia (Doan et al., 2017; Gould et al., 2014; Yu et al., 2013).

Classification studies allow the research of an optimized combination of multiple features for discriminating schizophrenia patients from healthy volunteers. Functionally, resting-state networks have been proven useful for classifying schizophrenia patients and controls with a high overall accuracy in independent training and testing data sets (Skatun et al., 2017). Support vector machine (SVM) have been proved that can classify small sample data of first-episode psychosis very well (Squarcina et al., 2017). In schizophrenia and autism spectrum disorder, classification studies have provided an unprecedented opportunity to improve the individualized diagnosis by means of radiomic signatures for mental disorders (Chaddad, Desrosiers, Hassan, & Tanougast, 2017; Cui et al., 2018). Emerging structural MRI studies further add empirical support to advance our understanding of accurate classification of schizophrenia (Liang et al., 2018; J. Liu et al., 2017; Qureshi, Oh, Cho, Jo, & Lee, 2017; Rozycki et al., 2018; Winterburn et al., 2017). More importantly, neuroanatomical subtypes of schizophrenia patients were linked with symptoms using machine learning techniques (Dwyer et al., 2018), aiding disease discrimination for biologically based diagnosis (Wang, Yu, Zhu, Yin, & Cui, 2019). The classification study combined with a multivariate method is a reliable option to explore the determinants of cortical neuroanatomical profile as defining features for schizophrenia.

Previous studies either examined imaging differences between patients with schizophrenia and controls, or used classification methods to identify features that could distinguish patients from controls. We hypothesized that neither all brain regions with differences could be used for classification, nor all classification features could have differences in schizophrenia. In this research, we used a multivariate computational approach that combined cortical features, including cortical thickness, surface area, gray matter volume, sulcal depth, metric distortion, and mean curvature, to investigate the abnormal changes between schizophrenia patients and HCs. In addition, we expected to observe multidimensional neuroanatomical patterns in classifying schizophrenia patients and HCs. Furthermore, we aimed to develop and validate a method of disease definition for schizophrenia by neuroanatomical features and explored whether these cortical features have equal contribution when differentiating the two groups, thus improving objective individualized schizophrenia identification using a quantitative and specific signature in clinical practice.

## 2. Methods

### 2.1. Subjects

The data set in this study included 52 first-episode schizophrenia patients at Xijing Hospital (51 inpatients and 1 outpatients without receiving any medication), and 66 HCs who were recruited by advertisement from the local community (Supplementary Table 1) (Cui, et al., 2018). There were a few overlapping subjects used in our previous study (Cui, Cai, et al., 2019; Cui et al., 2015; Cui, Wang, et al., 2017), but different MRI data were analyzed. Briefly, the exclusion criteria were comprised of: i) pregnancy, major medical and neurological diseases, history of significant head trauma, and illicit drug or alcohol abuse or dependence. ii) additional exclusion criteria for HCs included current or past history of psychiatric illness and the presence of psychosis in first-degree relatives. The absence of any psychotic syndromes in HCs was confirmed using the Prodromal Questionnaire (Loewy, Bearden, Johnson, Raine, & Cannon, 2005). Two senior clinical psychiatrists performed the neuropsychological assessments and diagnosis according to the Structured Clinical Interview for Diagnostic and Statistical Manual of Mental Disorders, Fourth Edition, Text Revision (DSM-IV-TR), as well as detailed information regarding past symptoms acquired through patient interview and examination of the patient’s medical records. Patients were assessed with the Positive And Negative Syndrome Scale (PANSS) on the day of scanning (Kay, Fiszbein, & Opfer, 1987). This study was approved by the local Research Ethics Committee (Xijing Hospital, Fourth Military Medical University). All participants gave written informed consent after a complete description of this study.

### 2.2. Data acquisition

The high-resolution, T1-weighted, three-dimensional (3D) anatomical data were performed in the Department of Radiology by using a 3.0-T Magnetom Trio Tim imaging unit with an eight-channel phased-array head coil (Siemens, Erlangen, Germany) and protocols published previously (Chang et al., 2017; Chang et al., 2015; Cui et al., 2016; Cui, Liu, et al., 2017; Cui, et al., 2018; B. Li et al., 2017; L. Liu et al., 2019). Imaging was acquired by using a magnetization-prepared rapid gradient-echo sequence with the following parameters: repetition time (TR) = 2530 ms, echo time (TE) = 3.5 ms, flip angle = 7°, field of view = 256 mm × 256 mm, data matrix = 256 × 256, section thickness = 1 mm, section gap = 0 mm, number of sections = 192, and image resolution = 1 × 1 × 1 mm.

### 2.3. Data processing

T1 Sequence image processing was performed using the Freesurfer image analysis suite (version 6.0.0, http://surfer.nmr.mgh.harvard.edu/). Briefly, preprocessing was performed with the following steps: i) skull stripping, ii) normalization to a standard anatomical template (Tournoux & Pierre, 1988), iii) correction for bias-field homogeneity, iv) segmentation of subcortical white matter and deep gray matter volumetric structures (Fischl et al., 2002; Fischl et al., 2004), v) gray-white mater boundary tessellation and a series of deformation procedures which consist of surface inflation (Dale, Fischl, & Sereno, 1999), vi) registration to a spherical atlas (Fischl, Sereno, & Dale, 1999) and parcellation of the cerebral cortex into units based on the gyral and sulcal structures (Fischl, et al., 2004). The information collected from preprocessing was used for calculating 408 structural measures, including volumetric (68 measures of cortical thickness, surface area and gray matter regional volume) and geometric (68 measures of mean curvature, metric distortion and sulcal depth) based on Desikan-Killiany Atlas (S. Li, et al., 2014). We calculated these six imaging measures of 68 regions parcellated according to the atlas, resulting in a total of 408 features (The definition for calculating the cortical features are shown in Supplementary Table 2). The data that the factors of sex and age of all subjects were regressive using FSL toolbox were reserved for further analyzing. Furthermore, we calculated and compared the group differences in the intracranial volume, brain volume and gray matter volume between patients and HCs to avoid their effects on cortical features (Supplementary Table 3).

### 2.4. Statistical analysis

Statistical analysis was mainly conducted by using Statistical Product and Service Solutions (SPSS) and R language (https://www.r-project.org/). Workflow proceeded as follows:

i. *t*-test: for each of the features, the independent two-tailed *t*-test was used to assess the differences between schizophrenia patients and HCs, while the threshold level in all features for significance criterion was determined at *P* < 0.05, uncorrected.
ii. Dimension reduction: 408 features (after removed the features which did not meet the condition: mean ± 2 * SD) were used in the classification, and the dimensionality of the features exceeded the sample size, creating difficulty in classification. Least absolute shrinkage and selection operator (LASSO) regression was used to achieve dimension reduction (Collins, Reitsma, Altman, & Moons, 2015; Huang et al., 2016). Due to the small sample size of our research and avoiding over-fitting, a leave-one-out cross validation approach was used in our research. We used leave-one-out cross validation to get a proper L1-regularization value (i.e., λ).

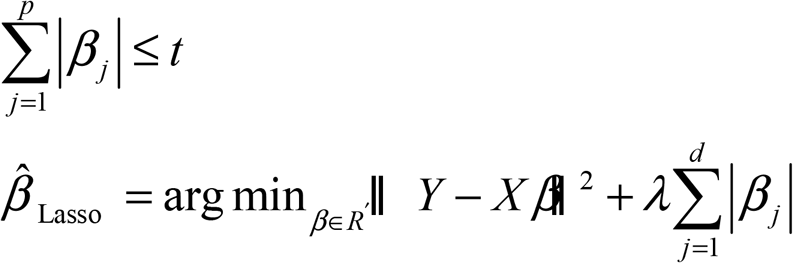 The exact relationship between λ and *t* is data-dependent. Tuning parameter (λ) selection in the LASSO model used leave-one-out cross validation via minimum criteria and the selected λ minimized the loss function. For each λ value and in each leave-one-out cycle, one subject was removed from the whole group and used as the test sample. The remaining subjects were used to train the model and the model was used to test the subject who had been removed. This procedure was repeated 1000 times. The misclassification was measured by the proportion of observations that patients were incorrectly classified into control groups for each λ value. The λ with the lowest misclassification was selected as the final λ. The remaining features were obtained. Finally, the different λ were trained using each individual feature. The dimension reduction algorithm used was glmnet (https://cran.r-project.org/web/packages/glmnet).
iii. Classification: the remaining features were used to build classification models using a SVM. In addition, the penalty parameter C of the error term was fixed at C=1 for all cases (default value). The classifier and the classifier was then used to test the subject who had been removed; the classification accuracy, specificity and sensitivity were measured and reported.
iv. Clinical correlation: the research index score was selected as the classification indicator to relate with the PANSS score. We integrated all of the classification indicators of each ROI after the dimension was reduced for each cortical feature.

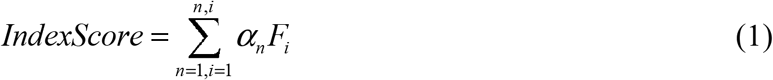

*α* : the classification indicator of each feature; F: cortical features; n: the number of ROIs after dimensionality reduction; and i: the number of cortical measures (i = 6).

The workflow is shown in Fig. 1.

**Fig. 1.**
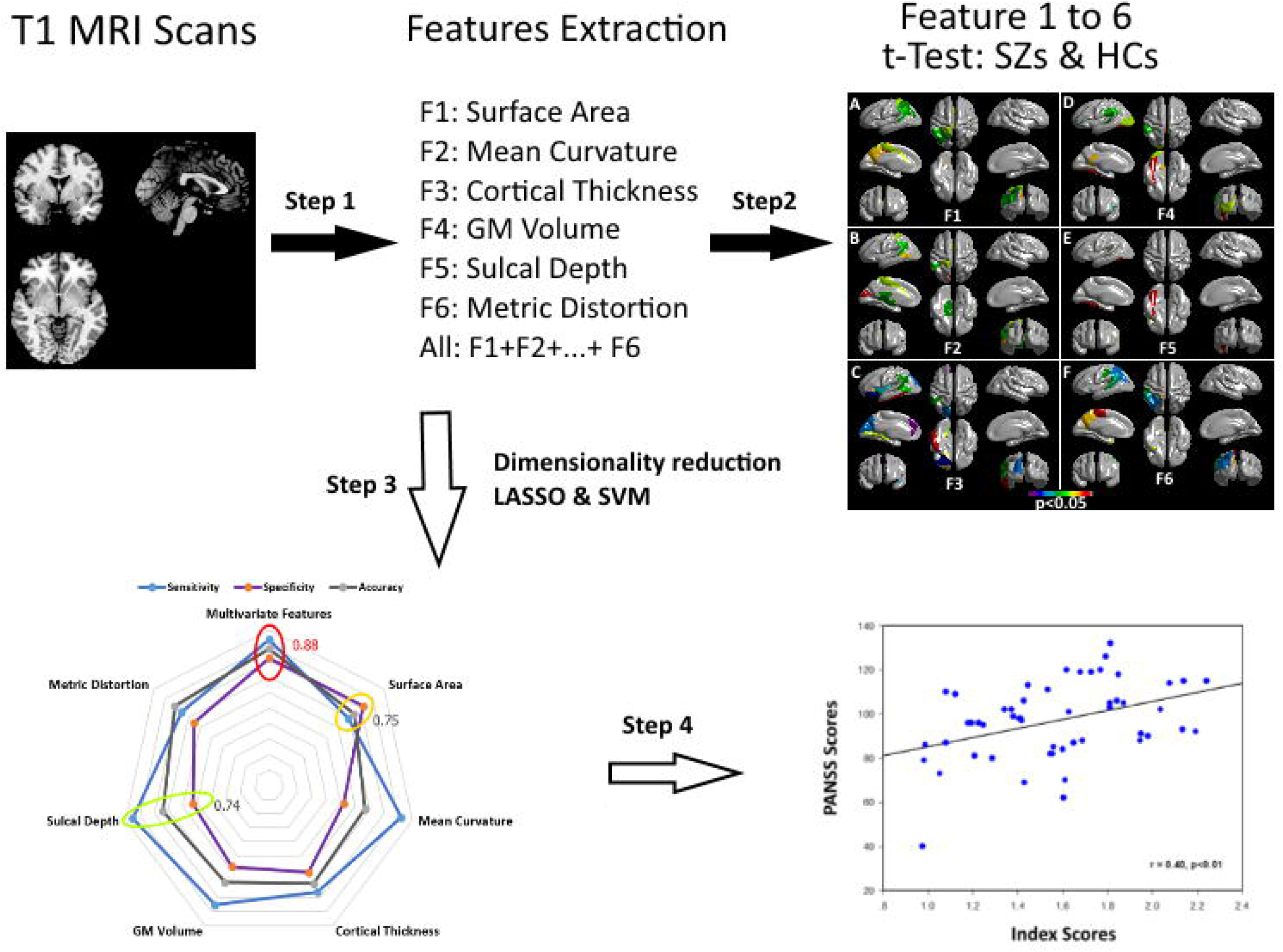
The workflow of data processing. Step 1: preprocessing and feature extraction of T1 MRI data; Step 2: the difference comparing SZs and HCs in each of the cortical features using a *t*-Test; Step 3: dimensionality reduction of all features in Step 1 using LASSO method and SVM algorithm; Step 4: correlation between index scores and PANSS scores.

### 2.5. Independent validation

We used an independent validation dataset (see the Supplement), and performed validation with features after dimensionality reduction in the principal dataset, as listed in Table 2.

**Table 1.**
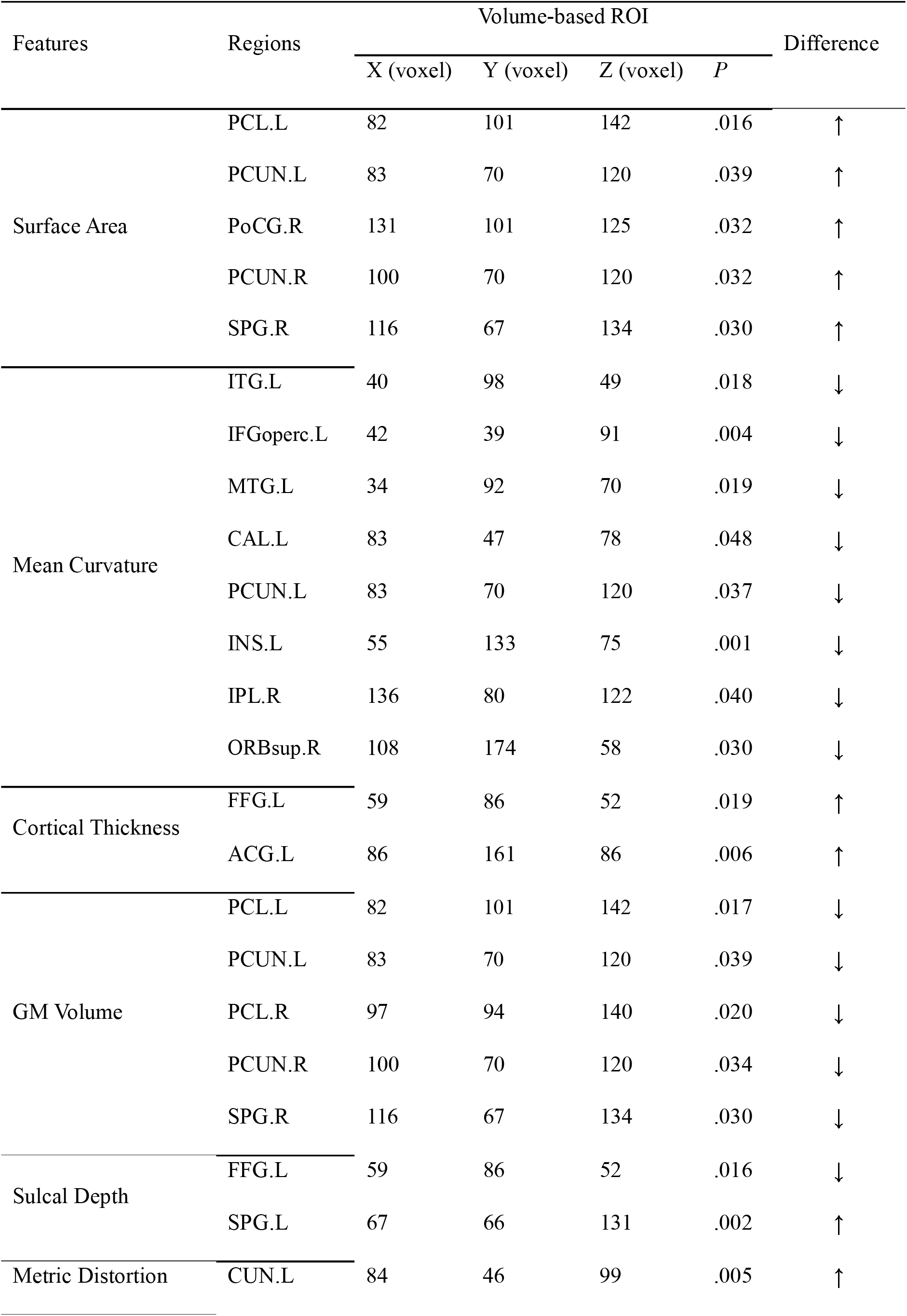
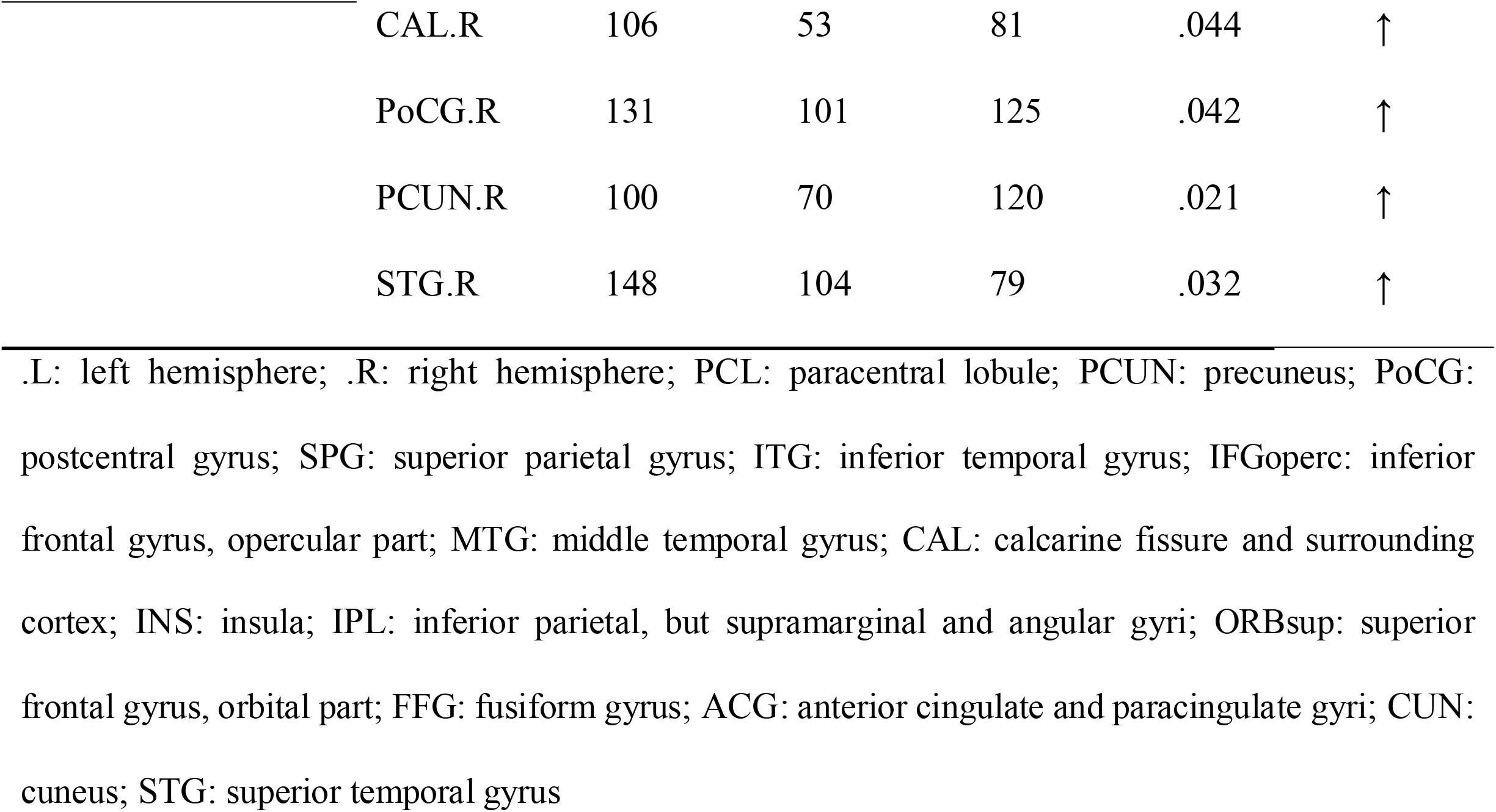
Each feature with significant differences between SZs and HCs.

**Table 2.** The features using the least absolute shrinkage and selection operator (LASSO) binary logistic regression model.

| Features | Regions | Weighted vector |
| --- | --- | --- |
| Mean Curvature | INS.L | 0.59 |
|  | ACG.L | 0.55 |
|  | IFGoperc.L | 0.22 |
|  | ACG.L | 0.01 |
| Cortical Thickness | FFG.L | 0.61 |
|  | ACG.R | 0.63 |
| GM Volume | EC.R | 0.07 |
|  | PCG.R | 0.43 |
| Sulcal Depth | PCG.L | 0.81 |
|  | IPL.L | 0.96 |
|  | LOG.L | 0.66 |
| Metric Distortion | CUN.L | 0.58 |
|  | STG.R | 0.85 |

L: left hemisphere, R: right hemisphere. INS: Insula, ACG: Anterior cingulate and paracingulate gyri, IFGoperc: Inferior frontal gyrus opercular part, FFG: Fusiform gyrus, EC: Entorhinal cortex, PCG: Posterior cingulate gyrus, IPL: Inferior parietal, LOG: lateral occipital gyrus, CUN: Cuneus, STG: Superior temporal gyrus.

## 3. Results

### 3.1. Multivariate computation-based abnormalities in patients

Abnormal areas in patients with schizophrenia are listed in Table 1 (*P* < 0.05, uncorrected for multiple comparisons, because our hypothesis indirectly concerned the difference of imaging). Apart from sulcal depth, other features showed a consistent trend in alterations, including increased surface area, cortical thickness, and metric distortion, as well as decreased mean curvature and gray matter volume. **Fig. 2** highlights these regions where *P* < 0.05 in each feature.

**Fig. 2.**
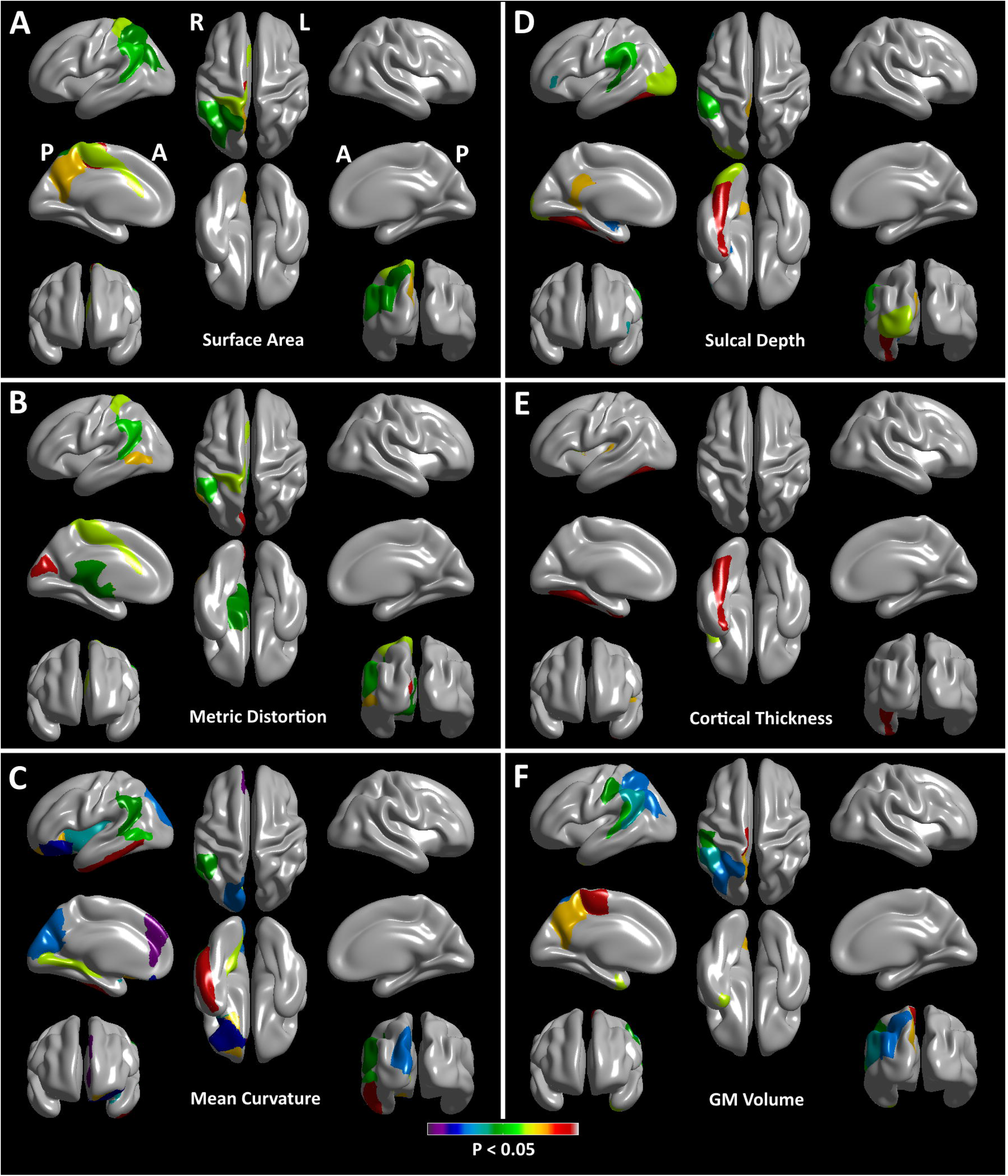
Differences in each of the cortical features. A: Surface Area; B: Metric Distortion; C: Mean Curvature; D: Sulcal Depth; E: Cortical Thickness; F: Grey Matter Volume. The regions where *P* values were less than 0.05 are colored.

### 3.2. Feature selection and classification

Of the cortical features, all features were reduced to 13 potential predictors with nonzero coefficients in the LASSO logistic regression model (Table 2). For the combination of six groups of imaging measures, the SVM classifier accurately discriminated patients from HCs on the basis of the ROC curve, with an accuracy of 88% (**Fig. 3** and Supplementary Fig. 1). The sensitivity and specificity were 94% and 82%, respectively. The classification parameters were repeated 1000 times and the average of each parameter were calculated. Specifically, after we find the optimal parameter, we get the dimension reduction feature, and without changing the parameter, we repeated the following SVM steps for 1000 times. As for each cortical feature, results are shown in Supplementary Table 4. Confirming these findings, similar results were observed in the independent validation (Supplementary Table 5).

**Fig. 3.**
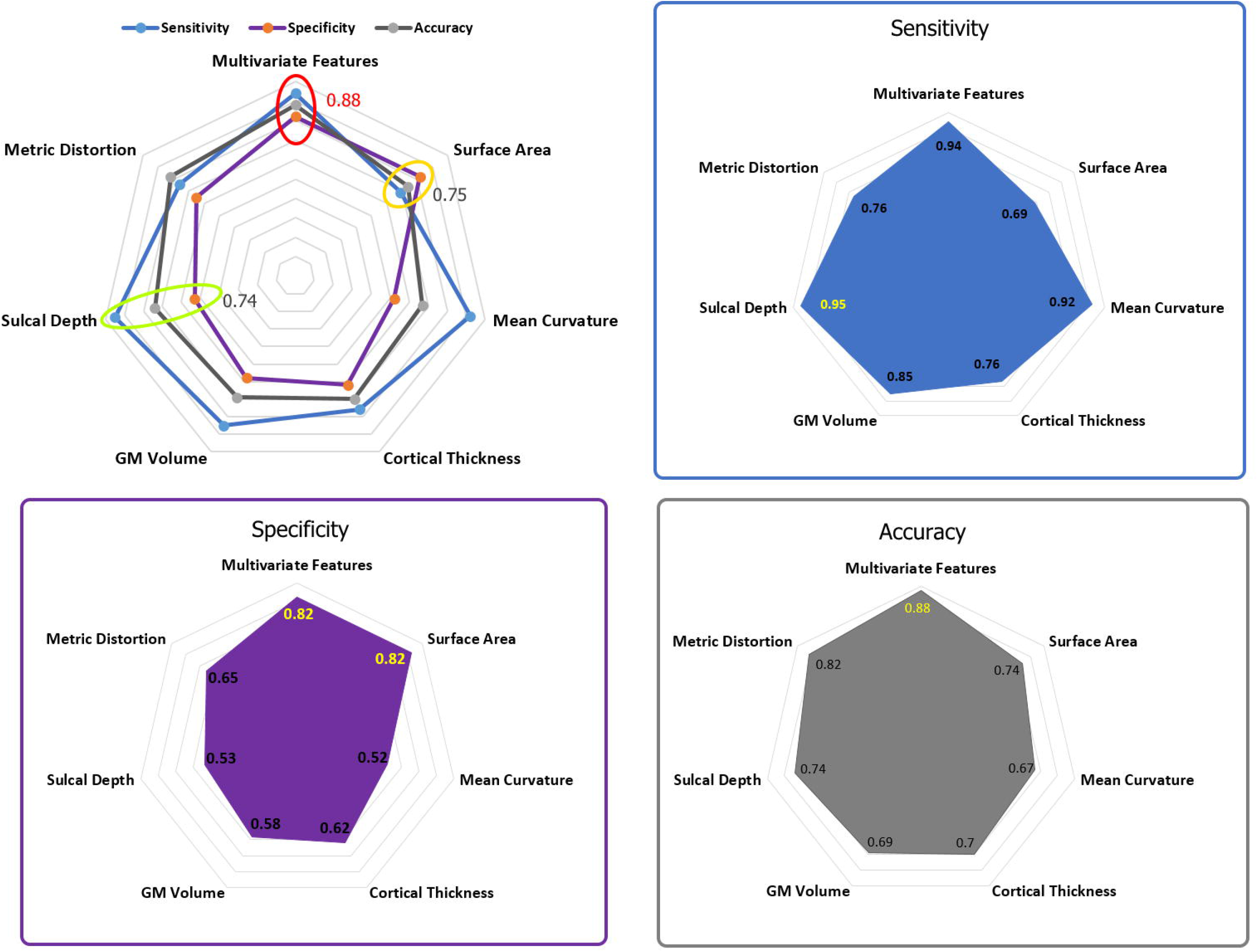
Classification of each of the cortical features and the combinations of the features.

### 3.3. Clinical correlates

In the correlation analysis, the index score derived from the multi-dimensional patterns was positively associated with the PANSS total score of patients (*r* = 0.40, *P* < 0.01).

## 4. Discussion

Based on cerebral cortical features in this study, we compared the differences between patients with schizophrenia and HCs using a multivariate computational method, and explored the diagnostic performance for schizophrenia via multi-dimensional neuroanatomical patterns. We found that not all abnormal brain regions are helpful for classification. Also, features used for classification do not necessarily have imaging measure differences. We detected the features of the mean curvature (left insula and left inferior frontal gyrus), cortical thickness (left fusiform gyrus), and metric distortion (left cuneus and right superior temporal gyrus) revealing both group differences and diagnostic capacity. The accuracy of identifying patients was 88% using optimized combination of all of the features, with a sensitivity of 94% and specificity of 82%.

The patterns of brain morphology in patients with schizophrenia that may be implicated in the pathophysiology are tangled with convergent findings based on the structural analysis. At the level of individual study, emerging evidence has shown dramatically varied neuroanatomical differences. First, as mounted by burgeoning studies, decreased volume of the anterior cingulate cortex with lower variability is one of the main findings of altered gray matter in schizophrenia (Brugger & Howes, 2017). Second, there has been a broad interest in cortical thickness in schizophrenia.

Third, reviewing literature for the past decade, increased parahippocampal-lingual and visual cortical gyrification (mean curvature) was detected in schizophrenia (Schultz et al., 2010; Schultz, et al., 2013), and mean curvature in the prefrontal cortex was related to integrity in short-range cortico-cortical connections and clinical outcome in schizophrenia (Lubeiro et al., 2017). Fourth, other cerebral morphological features, metric distortion and sulcal depth, were also compared between schizophrenic and healthy populations. Patients showed markedly reduced metric distortion in the Broca’s area (pars triangularis of the left inferior frontal gyrus) in the patient group relative to the controls (Wisco, et al., 2007). They exhibited shallower olfactory sulci (Turetsky, Crutchley, Walker, Gur, & Moberg, 2009), and there was a significant association between neuroleptic exposure and depth of the right paracingulate sulcus (Rametti et al., 2010). At present, we demonstrated a new perspective of morphometric abnormalities in schizophrenia in line with biological heterogeneity demonstrated by a recently published meta-analysis (Brugger & Howes, 2017). Nevertheless, part of this substantial evidence is the fundamental basis for defining schizophrenia via cortical features in nature and extent.

Of primary importance is that this study extends previous results in two aspects. Our study not only validates that there are significant differences in cortical characteristics between schizophrenia patients and healthy population, but also finds that the characteristics of imaging differences are not identical to the results of dimensionality reduction by machine learning. On one hand, the accuracy was between 73.0% and 87.09% in classifying schizophrenia by functional connectivity or resting-state network features in previous (Anderson et al., 2010; Ariana & Cohen, 2013; Jafri, Pearlson, Stevens, & Calhoun, 2008; Shen, Wang, Liu, & Hu, 2010) and recent machine learning studies (Cui, et al., 2018; Cui, Wei, et al., 2019; Mikolas et al., 2016; Skatun, et al., 2017). We utilized cortical features to obtain an accuracy of 88% in the present study, increasing the diagnostic performance of neuroimaging in schizophrenia identification and pushing preclinical findings up to a level of clinical translation. On the other hand, we shared the same sample in the current and previous studies (Cui, et al., 2018), since we planned to further investigate the practical value of structural neuroradiology in schizophrenia. As a result, both multi-dimensional neuroanatomical patterns and functional connectivity features hold great promise for assisting diagnosing schizophrenia. Multi-dimensional neuroanatomical pattern analysis is another reliable option for classification study in schizophrenia.

Another important implication of our results is that we provided new insights into the determinants of differences of the underlying neurobiology and unaltered structure functioning as diagnostic tools by means of a multivariate computational approach and multidimensional neuroanatomical patterns, with implications for precision medicine. Generally speaking, multiple lines of evidence have characterized schizophrenia patients via certain abnormalities in neuroanatomy, and of particular interest is cortical or subcortical gray matter, through different methods. These findings are supportive of generalizability across heterogeneous samples (Dietsche, Kircher, & Falkenberg, 2017). In our study, there were 408 features that were dimensionally reduced by LASSO and classified by SVM. The results show that not all the differences of imaging between patients with schizophrenia and controls are the characteristics of distinguishing patients and controls, vice versa. Through independent verification of another data set, we believe that the features after dimensionality reduction could be used to identify patients with schizophrenia accurately.

As compared with previous literatures by Yu et al, Gould et al, and Doan et al who also used multivariate approach for classification of patients with schizophrenia and HCs (Doan, et al., 2017; Gould, et al., 2014; Yu, et al., 2013), the current study proposes a point of view, i.e., neither all brain regions with differences could be used for classification, nor all classification features could have differences in schizophrenia. In the study by Doan et al, they used multivariate machine learning analysis based on cortical thickness, surface area, and gray matter density maps. They found six biologically meaningful patterns showing group effects sensitive to schizophrenia (Doan, et al., 2017). In our study, for the features after LASSO dimensionality reduction, we provided classification weights (the contribution degree in the process of classification). Additionally, the features we used included the features as selected by Doan et al and several other cortical features, suggesting the best levels of accuracy, specificity and sensitivity for the integrated features. Similar results were found in the independent validation. Gould et al used SVM to classify patients and controls based on the volume of gray matter and white matter, showing a cross-validation accuracy of 71%. They also discussed the effects of cognitive subtypes and gender alignment rates (Gould, et al., 2014). In addition to features of volume, we also examined other five features calculated. However, cortical volume was not the most prominent feature for classification in our study, with an classification of 69% (Supplementary Table 4). Another study with a relatively small sample size based on SVM using functional connectivity, achieving 62.0% accuracy for identifying schizophrenia via leave-one-out cross validation (Yu, et al., 2013). Instead of functional patterns, structural neuroimaging was performed in this present study. Of note, independent validation is helpful to enhance the generalization of our results.

Despite these encouraging results, there is a drawback in the current study. Information about smoking was missing for a portion of participants, and alcoholic consumption was unavailable. How much the contribution of these factors is unknown. Additionally, structural imaging is a promising tool of predicting schizophrenia patients’ subsequent response to antipsychotics (Altamura et al., 2017; Dusi et al., 2017; Morch-Johnsen et al., 2015); however, this study is limited on the issue of diagnosis. Also prognostic significance of the cortical features via structural MRI is of importance for schizophrenia in clinical settings. Biologically-based prediction of the treatment response in schizophrenia seems to be urgent, because clinical decision-making requires the guidance of quantitative and objective tests.

In summary, linking abnormalities to disease diagnosis can form a valuable method for managing schizophrenia using multivariate computation and multi-dimensional patterns via high-resolution structural MRI. This study detects widespread cortical abnormalities in schizophrenia and develops a multi-dimensional neuroanatomical patterns-based approach to classify schizophrenia. The cortical features might be a promising tool for correct identification of patients with schizophrenia.

## Data Availability

The datasets generated for this study are available on request to the corresponding author.

## Contributors

L.L. and L.-B.C. contributed equally to this work. Guarantors of integrity of entire study, L.B.C., H.Y., W.Q.; study concepts/study design or data acquisition or data analysis/interpretation, all authors; manuscript drafting or manuscript revision for important intellectual content, all authors; approval of final version of submitted manuscript, all authors; agrees to ensure any questions related to the work are appropriately resolved, all authors; literature research, L.-B.C., X.-S.W., Y.-B.X., H.Y.; clinical studies, L.L., L.-B.C., D.W., S.Q., Y.-H.Z., H.-N.W., H.Y.; experimental studies, L.-B.C., Y.-B.X., P.H., H.Y.; statistical analysis, L.L., L.-B.C., N.-B.F., Z.-L.X., H.Y., W.Q.; and manuscript editing, L.L., L.-B.C., K.M.von K., H.Y., W.Q.

## Acknowledgements

We are indebted to Chen Li, the Department of Radiology, Xijing Hospital, Fourth Military Medical University, for his helpful suggestions on clinical data acquisition.

